# Clinical Characteristics and Outcomes of Cholera Cases in Lusaka and Eastern Provinces of Zambia, January 2023 to July 2024 Cholera Outbreaks: A Retrospective Facility-Based Study

**DOI:** 10.1101/2025.11.05.25339557

**Authors:** Chipo Nkwemu, Nedah Chikonde Musonda, Samson Shumba, Deborah Tembo, Miyanda Simwaka

## Abstract

Cholera remains one of the major public health challenges in sub-Saharan Africa due to poor water, sanitation, and hygiene (WASH) systems and climate change (floods, droughts). Zambia had major outbreaks between 2023 and 2024, yet there is limited information about clinical characteristics and outcome predictors for the patients who were affected. Here, we describe clinical profiles and outcomes of cholera patients while identifying factors associated with mortality.

A retrospective cross-sectional study design was conducted using secondary data from the national cholera surveillance system. The study included those with confirmed cholera cases documented in Lusaka and Eastern Provinces from January 2023 to July 2024. Patients with incomplete records were excluded. Descriptive statistics, bivariate tests, and multivariable logistic regression were used to assess clinical characteristics, outcomes, and factors associated with mortality. A total of 2,844 confirmed cases were analyzed, with 97% from Lusaka and 3% from Eastern Province. The median age was 24 years (IQR: 6–35), with Eastern Province patients being older (median 34 years). Males comprised 53.3% of cases. Most patients (99.6%) were admitted as inpatients, with a mean hospital stay of 1.8 days. Overall, 95.6% survived and 4.4% (n=126) died. Mortality was highest among patients ≥65 years (17.1%) and infants <1 year (8.5%). Male patients (5.9% vs. 2.7% in females, p<0.001) and outpatients (54.8% vs. 3.9% in inpatients, p<0.001) were at greater risk. Multivariable analysis showed increased odds of death among those aged 25–44 (AOR 2.16, 95% CI: 1.25–3.73) and ≥65 years (AOR 5.34, 95% CI: 2.62–10.87). Being female (AOR 0.42, 95% CI: 0.27–0.63) and inpatient care (AOR, 95% CI: 0.02–0.07) were protective.

Adults 65 years and above, infants, males, and outpatients experienced the highest mortality during the outbreaks. Inpatient care was very protective, which shows how important it is to find high-risk patients early, refer them quickly, and admit them into care. To reduce the number of cholera-related deaths in Zambia, it is important to strengthen triage systems, teach people in the community, do WASH interventions, and improve preparedness.

## Background

Cholera is still a major public health threat globally, with the seventh pandemic resurging recently. It is an acute, often rapidly fatal diarrheal disease that can kill within hours if untreated ^[1]^. *Vibrio cholerae* is a gram-negative, motile bacterium that thrives in brackish water and estuarine environments ^[2]^. Two serogroups, O1 and O139, are responsible for epidemic and pandemic cholera ^[1–3]^. The bacterium produces cholera toxins that attach to GM1 ganglioside receptors on enterocytes, raise the levels of cyclic AMP inside cells, and cause a huge amount of water and electrolytes into the intestinal lumen ^[4]^. Cholera is transmitted mainly through the fecal-oral route, primarily through the ingestion of water or food contaminated with *Vibrio cholerae* from fecal sources. Secondary person-to-person transmission may also occur in environments characterized by poor sanitation and hygienic practices ^[4, 5].^ Improvement in water, sanitation, and hygiene (WASH), oral cholera vaccination, health education, and strong surveillance systems are very important when it comes to cholera prevention ^[6]^ [6]. Coordinated interventions have also been shown to reduce incidence during outbreaks, yet infrastructure gaps continue to fuel endemic transmission in parts of Africa and Asia ^[4–6]^ Some of the contributing elements to Vibrio cholerae proliferation are environmental factors such as poor sewage treatment and open defecation, contaminated surface and groundwater, and climate events (e.g., flooding, drought) that compromise water infrastructure ^[7]^.

According to the WHO situation report, December 18th, 2024, a total of 733,956 cholera cases and 5,162 cholera-related deaths were reported between January and November 2024 in 33 different countries in five WHO regions ^[8]^. Between January 2024 and March 2025, approximately 178,000 cholera cases and 2,900 deaths recorded in Eastern and Southern Africa were associated with limited access to safe water, sanitation, hygiene, and health services ^[7, 9].^ Zambia declared cholera outbreak in October 2023, and this moved into a critical public health emergency with 14,900 cases reported and 560 deaths (3.8% fatality rate) ^[10]^. Government response measures included the establishment of cholera treatment centers, oral rehydration points, and vaccination campaigns in the hotspot areas around the country, including Lusaka, to limit spread. Despite these efforts, some cases were still noted in different parts of the country notably in Lusaka and eastern provinces, reflecting challenges in water, sanitation, and hygiene infrastructure ^[11]^.

Although national surveillance provides epidemiological trends, detailed data on clinical presentations, treatment outcomes, and risk factors among Zambian cholera patients remain scarce. There is also a lack of detailed, facility-based data describing the clinical characteristics and outcomes of cholera cases in Lusaka and Eastern provinces. Without such information, it is difficult to identify context-specific risk factors for poor outcomes and improve clinical management protocols. Additionally, published literature from Zambia has mainly looked at outbreak responses strategies and water, sanitation, and hygiene (WASH) intervention, with little attention on clinical profiles and outcomes of patients managed during the recent outbreaks

This study aims to characterize the clinical features and the outcomes of cholera cases managed in health facilities during the January 2023 to July 2024 outbreaks in Lusaka and Eastern Provinces of Zambia. By providing facility-based evidence, we seek to bridge the gap and inform local and national cholera response strategies, improve clinical care, and reduce cholera-related mortality.

### Conceptual framework for Clinical outcomes of Cholera

This framework adopts and adapts determinants identified in global, regional, and national reports to illustrate how upstream factors translate into clinical outcomes, as shown in **Fig. 1** below.

**Fig. 1.**
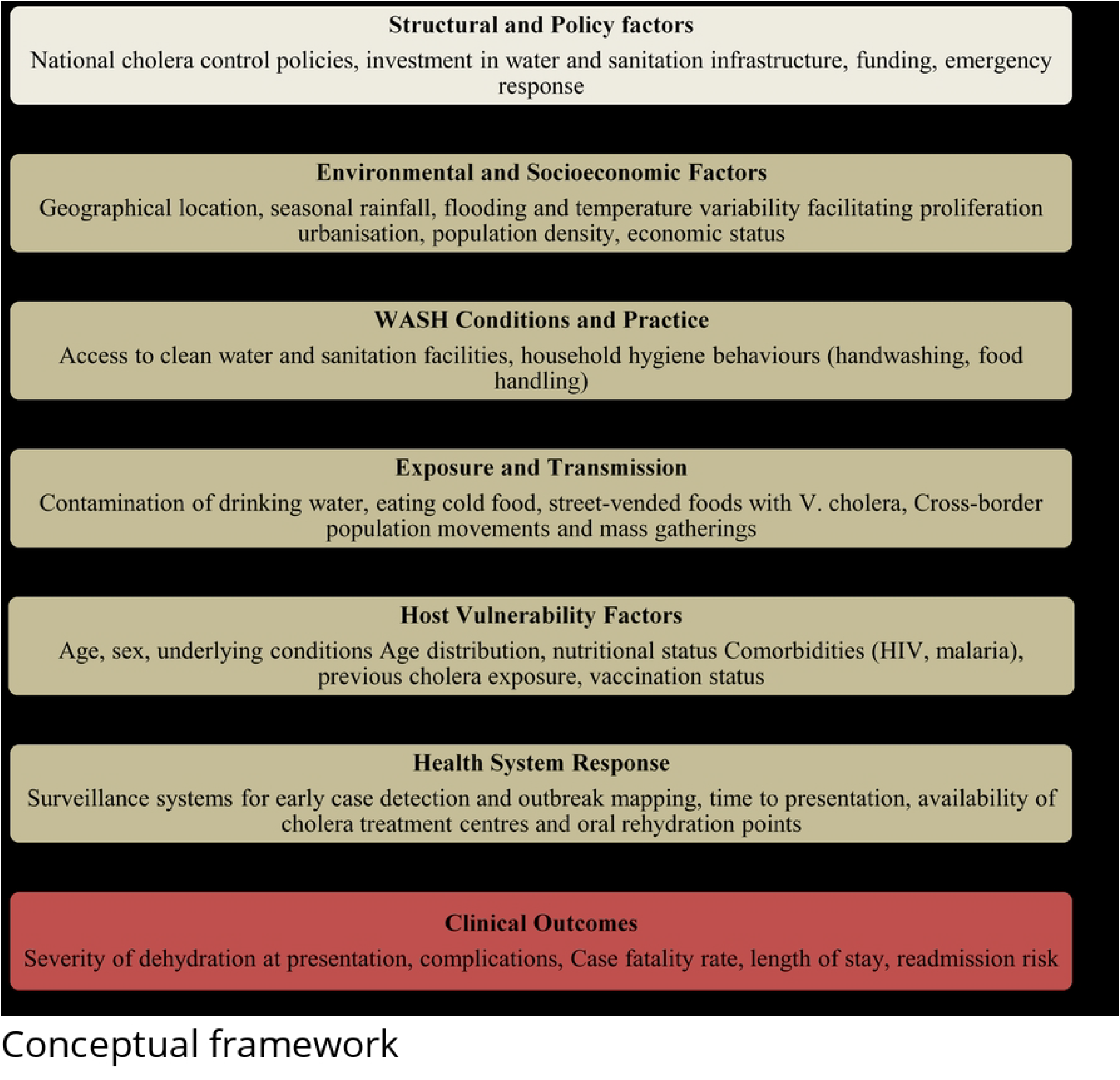
Conceptual framework.

## Methodology

### Study design

This was a retrospective cross-sectional study utilizing secondary data from the national cholera surveillance system during the 2023–2024 cholera outbreak in Zambia. The primary objective was to analyze the clinical characteristics and outcomes of cholera patients managed within the national healthcare system in Zambia.

### Setting and population

The study was conducted in the Lusaka and Eastern provinces of Zambia, with data aggregated from all health facilities across the two provinces that reported cholera cases. The study population included all suspected and confirmed cholera cases recorded between 1^st^ January 2023 and 31^st^ July 2024.

### Inclusion Criteria

1. All suspected or confirmed cholera cases registered within the specified time frame in the Lusaka and Eastern provinces of Zambia.
2. The study includes cases with complete records for the variables of interest.

### Exclusion criteria

1. Cases with incomplete or missing data for key variables were excluded from the final analysis to ensure data quality and reliability.

### Data sources and collection

Data was extracted from the national cholera surveillance system, which is managed by the Zambia Ministry of Health. Patient-level records were obtained from a combination of facility-based registers and the electronic cholera case reporting tools. The data extraction process involved systematic review and compilation of these records into a unified dataset for analysis.

### Data management and analysis

Data was checked to ensure completeness and accuracy. We conducted descriptive data analysis to show the trend of epidemic cholera admissions over time, as well as the mean, standard deviation, median, and interquartile range for continuous variables; additionally, we disaggregated the baseline characteristics by province. Bivariate analysis of factors associated with mortality in cholera patients and multivariable analysis, adjusting for age and sex, was conducted to further understand the factors associated with death among cholera patients. We analyzed data using Stata, version 19.0 (StataCorp LLC, College Station, Texas, USA) and R https://www.R-project.org/.

### Ethical considerations

We obtained ethical approval from the University of Zambia Biomedical Research Ethics Committee (UNZABREC **ref: No 5066-2024**). We also obtained permission to undertake the study from the Zambia National Public Health Institute (ZNPHI).

## Results

A total of 3,487 clients with suspected cholera cases were extracted from data from Eastern and Lusaka provinces, and 16.7% (581/3,487) with missing diagnosis results were excluded from the data. Of the 2,900 with confirmed diagnoses, 98.1% (2,844) had cholera, and 97.6% (2,718) survived while 4.4% (126) died **(Fig. 2)**.

**Fig. 2:**
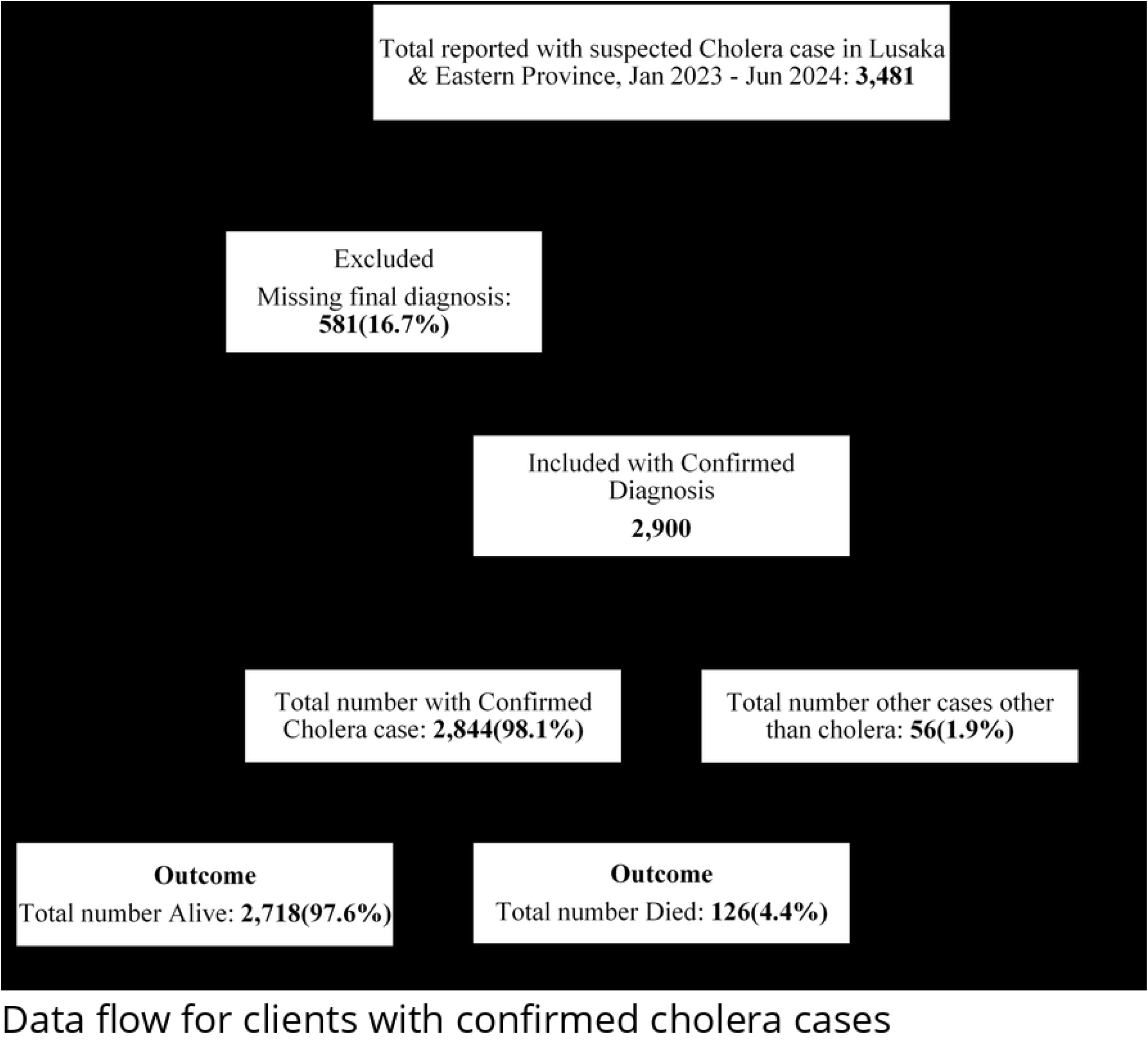
Data flow for clients with confirmed cholera cases

Fig. 3 below shows the cholera outbreaks that occurred between January 2023 and July 2024 and the distribution of cholera admissions in the Lusaka and Eastern Provinces of Zambia. The first outbreak was of a lower grade compared to the second outbreak, with the first small cluster of admissions, mainly of low-level activity, occurring in early 2023 and in late 2023, followed by an explosive peak in January 2024 at nearly 300 daily admissions. This peak was later followed by a gradual decline of cholera admissions over the period until July 2024.

**Fig. 3:**
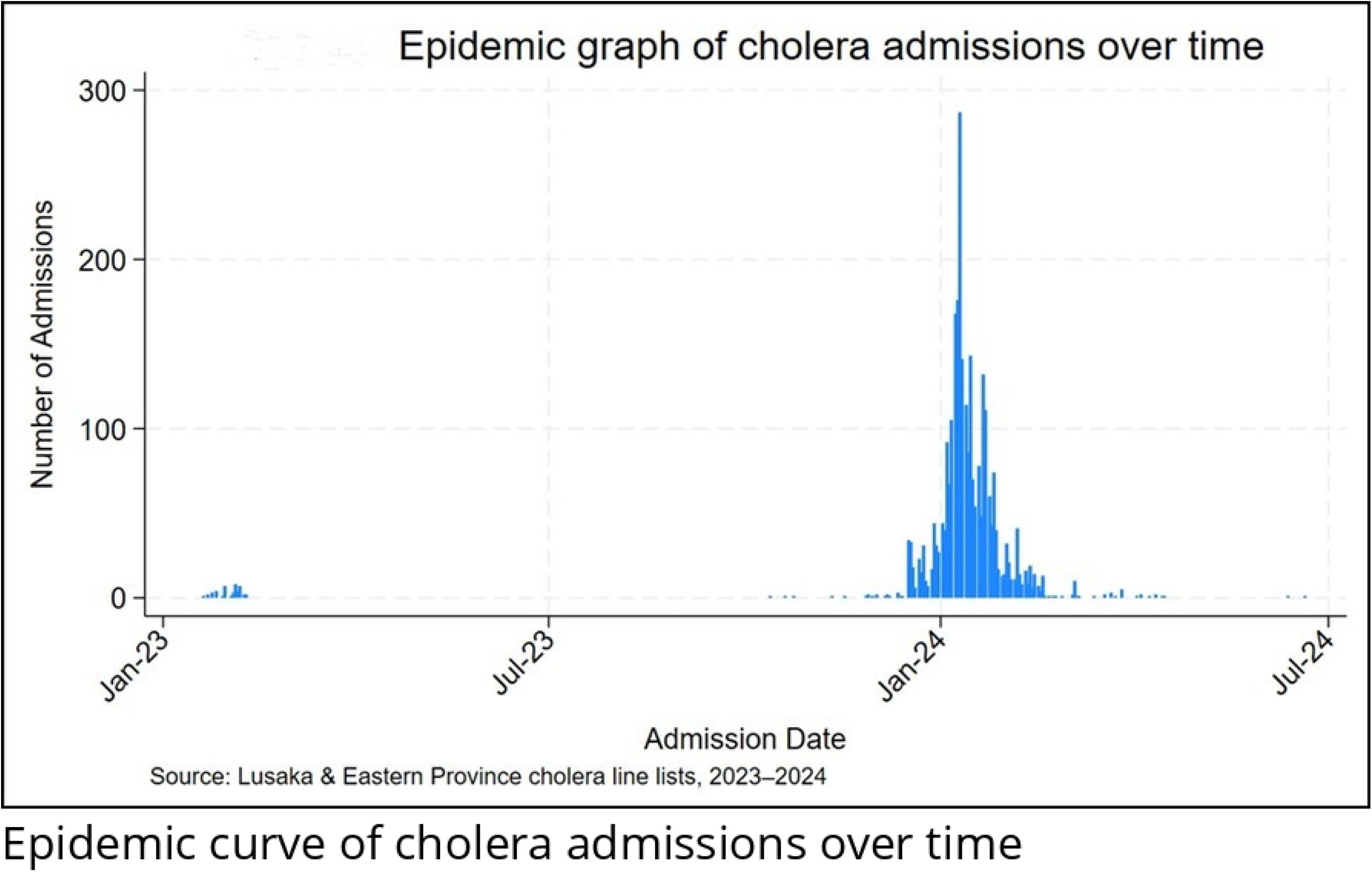
Epidemic curve of cholera admissions over time

A total of 2,844 cases were included in the study, with 97% (n=2,759) coming from Lusaka province and 3% (n=85) from Eastern Province. The overall median age was 24 years (IQR: 6-35), with patients from Eastern Province tending to be older (median: 34 years; IQR: 23-42) compared to those from Lusaka Province (median: 23 years; IQR: 6-35). The largest proportion of cases (34.8%, n=991) occurred in the 25-44-year age group. Children under one year of age accounted for 2.5% of cases (n=71), all of whom were from Lusaka Province. Slightly more than half of all the patients were males (53.3%, n=1517). Eastern Province had a higher proportion of males at 62.4% (n=52) compared to Lusaka with 53.1% (n=1,464). Almost all the patients were admitted as inpatients (99.6%, n=2,832), with the Eastern Province recording no outpatients. Rapid diagnostic testing for cholera was performed on 7.6% (n=215) of cases, more frequently in Eastern Province (23.5%, n=20) than in Lusaka (7.1%, n=190). Over half of the patients (54.8%, n=1559) had unknown test status.

The mean hospital stay was 1.8 days (SD: 0.8) overall, with a similar average in both provinces. Most patients stayed 0–1 day (40.9%, n=1162), though a greater proportion of Eastern Province patients stayed 2–3 days (69.4% vs. 38.3% in Lusaka). The mean delay from symptom onset to arrival at a health facility was 1.2 days overall, slightly longer in Eastern Province (1.4 days) compared to Lusaka Province. Three-quarters of patients (75.0%, n=2134) presented without any delay, with Eastern Province reporting a higher proportion of delayed presentations (45.9% vs. 24.3%) compared to Lusaka. Antibiotic use was uncommon (3.2%, n=92), but much higher in Eastern Province (34.1%, n=29) compared to Lusaka (2.3%, n=63). Overall, most patients had a favorable clinical outcome (95.6%, n=2718). Mortality rates were similar across provinces, with 4.4% in Lusaka compared to 4.7% in Eastern.

**Table 1:** Baseline Characteristics of Cholera Patients by Province (Lusaka and Eastern Provinces)

| Characteristics | Category | Total Cases<br>N=2844 | Province |  |
| --- | --- | --- | --- | --- |
|  |  |  | Lusaka<br>N = 2759<br>(97.0%) | Eastern<br>N = 85 (3.0%) |

| Age | Age in Years<br>[median (IQR)] | 24 (6–35) | 23 (6 – 35) | 34 (23–42) |
| --- | --- | --- | --- | --- |
| Age bands (years) | <1 | 71 (2.5) | 71 (2.6) | 0 (0.0) |
|  | 1-4 | 526 (18.5) | 523 (19.0) | 3 (3.5) |
|  | 5-14 | 425 (14.9) | 418 (15.2) | 7 (8.2) |
|  | 15-24 | 476 (16.7) | 458 (16.6) | 18 (21.2) |
|  | 25-44 | 991 (34.8) | 953 (34.5) | 38 (44.7) |
|  | 45-64 | 273 (9.6) | 257 (9.3) | 16 (18.8) |
|  | 65+ | 82 (2.9) | 79 (2.9) | 3 (3.5) |
| Gender | Male | 1,517 (53.3) | 1,464 (53.1) | 53 (62.4) |
|  | Female | 1,327 (46.7) | 1,295 (46.9) | 32 (37.6) |
| Patient category | Inpatient | 2,832 (99.6) | 2,747 (99.6) | 85 (100.0) |
|  | Outpatient | 12 (0.4) | 12 (0.4) | 0 (0.0) |
| Rapid diagnostic test (RDT) done | Yes | 215 (7.6) | 195 (7.1) | 20 (23.5) |
|  | No | 1,070 (37.6) | 1,005 (36.4) | 65 (76.5) |
|  | Unknown | 1,559 (54.8) | 1,559 (56.5) | 0 (0.0) |
| Days stayed at the Hospital | Days in Hospital<br>[Mean (SD)] | 1.8 (0.8) | 1.8 (0.8) | 1.8 (0.5) |
| Days stayed at the hospital | 0 to 1 day | 1,162 (40.9) | 1,142 (41.4) | 20 (23.5) |
|  | 2-3 days | 1,115 (39.2) | 1,056 (38.3) | 59 (69.4) |
|  | 4+ days | 567 (19.9) | 561 (20.3) | 6 (7.1) |
| Days delayed going to facility | Days delayed to HF<br>[Mean (SD)] | 1.2 (0.5) | 1.2 (0.4) | 1.4 (0.5) |
| Days delayed going to facility | 0 days | 2,134 (75.0) | 2,088 (75.7) | 46 (54.1) |
|  | 1+ days | 710 (25.0) | 671 (24.3) | 39 (45.9) |
| Antibiotic given | No | 2,752 (96.8) | 2,696 (97.7) | 56 (65.9) |
|  | Yes | 92 (3.2) | 63 (2.3) | 29 (34.1) |
| Unfavorable outcome | Survived | 2,718 (95.6) | 2,637 (95.6) | 81 (95.3) |
|  | Died | 126 (4.4) | 122 (4.4%) | 4 (4.7%) |

### Bivariate analysis

The median age of patients who died was significantly higher than that of survivors (32 years vs. 23 years, p<0.001). Mortality varied markedly across age groups (p<0.001), increasing from 1.2% among those aged 5–14 years to 17.1% among those aged ≥65 years. Infants (<1 year) also had a relatively high mortality rate (8.5%). Males, compared to females, had a higher mortality rate, 5.9% vs. 2.7%, p<0.001. Patient category was strongly associated with outcome, with outpatients experiencing much higher mortality than inpatients, 54.8% vs. 3.9%, p<0.001.

Rapid diagnostic testing was also linked to higher observed mortality (p<0.001): 9.8% among those tested, compared to 6.4% among those not tested, and 2.3% among those with unknown test status. The mean length of hospital stay was slightly shorter among those who died (1.8 vs. 2.2 days, p=0.046), and mortality differed across stay categories (p=0.014), being highest among those hospitalized for ≤1 day (5.8%) compared to those hospitalized for 2–3 days (3.7%) or ≥4 days (3.2%). Delay in presentation to a health facility was not significantly associated with mortality (mean delay 0.5 vs. 0.4 days, p=0.51). Similarly, neither antibiotic use (p=0.95) nor province (p=0.90) was associated with differences in mortality.

**Table 2:** Bivariate analysis of factors associated with mortality in cholera patients.

| Characteristics | Total Cases<br>N = 2,844 | Outcome |  | p-value |
| --- | --- | --- | --- | --- |
|  |  | Survived<br>n (%)<br>n = 2,718 | Died<br>n (%)<br>n = 126 |  |
| **Age in Years [median (IQR)]** | 24 | 23.0 (-) | 32.0 (-) | <0.001 |
| **Age bands (years)** |  |  |  | <0.001 |
| <1 | 71 | 65 (91.5) | 6 (8.5) |  |
| 1-4 | 526 | 505 (96.0) | 21 (4.0) |  |
| 5-14 | 425 | 420 (98.8) | 5 (1.2) |  |
| 15-24 | 476 | 463 (97.3) | 13 (2.7) |  |
| 25-44 | 991 | 949 (95.8) | 42 (4.2) |  |
| 45-64 | 273 | 248 (90.8) | 25 (9.2) |  |
| 65+ | 82 | 68 (82.9) | 14 (17.1) |  |
| **Gender** |  |  |  | <0.001 |
| Male | 1,517 | 1,427 (94.1) | 90 (5.9) |  |
| Female | 1,327 | 1,291 (97.3) | 36 (2.7) |  |
| **Patient category** |  |  |  | <0.001 |
| Inpatient | 2,813 | 2,704 (96.1) | 109 (3.9) |  |
| Outpatient | 31 | 14 (45.2) | 17 (54.8) |  |
| **A rapid test is done.** |  |  |  | <0.001 |
| Yes | 214 | 194 (90.2) | 21 (9.8) |  |
| No | 1,027 | 1,001 (93.6) | 69 (6.4) |  |
| Unknown | 1,603 | 1,523 (97.7) | 36 (2.3) |  |
| **Days stayed at the Hospital [Mean (SD)]** | 2.1 | 2.2 (1.8) | 1.8 (1.8) | 0.046 |
| **Days stayed at the hospital** |  |  |  | 0.014 |
| 0-1 day | 1162 | 1,095 (94.2) | 67 (5.8) |  |
| 2-3 days | 1115 | 1,074 (96.3) | 41 (3.7) |  |
| 4+ days | 567 | 549 (96.8) | 18 (3.2) |  |
| **Days delayed to Hospital [Mean (SD)]** | 0.4 | 0.4 (0.9) | 0.5 (1.1) | 0.51 |
| **Days delayed to Hospital** |  |  |  | 0.35 |
| 0 days | 2,134 | 2,035 (95.4) | 99 (4.6) |  |
| 1+ days | 710 | 683 (96.2) | 27 (3.8) |  |
| **Antibiotic given** |  |  |  | 0.95 |
| Yes | 2,751 | 2,629 (95.6) | 122 (4.4) |  |
| No | 93 | 89 (95.7) | 4 (4.3) |  |
| **Province (Location)** |  |  |  | 0.90 |
| Lusaka | 2,759 | 2,637 (95.6) | 122 (4.4) |  |
| Eastern | 85 | 81 (95.3) | 4 (4.7) |  |

### Multivariable Logistic Regression of Predictors of Death

In the multivariable analysis, adjusting for age and sex, several factors were significantly associated with mortality among cholera patients. Compared with patients aged 45–64 years, those aged 5–14 years had significantly lower odds of death (AOR = 0.31, 95% CI: 0.12–0.80), while those aged 25–44 years (AOR = 2.16, 95% CI: 1.25–3.73) and ≥65 years (AOR = 5.34, 95% CI: 2.62–10.87) had significantly higher odds of death. Female patients had 58% lower odds of death compared to males (AOR = 0.42, 95% CI: 0.27– 0.63). Being admitted as an inpatient was strongly protective (AOR = 0.04, 95% CI: 0.02– 0.07) compared to being treated as an outpatient. Length of hospital stay was also associated with mortality. Compared with patients hospitalized for 0–1 day, those hospitalized for 2–3 days (AOR = 0.53, 95% CI: 0.34–0.80) or ≥4 days (AOR = 0.48, 95% CI: 0.28–0.85) had significantly reduced odds of death. Province of treatment, rapid diagnostic testing, delay in presentation, and antibiotic use were not significantly associated with mortality after adjustment.

**Table 3:** Multivariable Logistic Regression of Predictors of Death.

| Characteristics | Category | Factors associated with Death |  |
| --- | --- | --- | --- |
|  |  | Crude analysis<br>OR (95% CI) | Multivariable analysis<br>AOR [95% CI] |
| **Age bands (years)** | 45-64 | Ref | Ref |
|  | <1 | 2.09 (0.86, 5.09) | 2.03 (0.81, 5.09) |
|  | 1-4 | 0.94 (0.55, 1.60) | 1.07 (0.62, 1.85) |
|  | 5-14 | 0.27 (0.11, 0.68) | 0.31 (0.12, 0.80) |
|  | 15-24 | 0.63 (0.34, 1.19) | 0.70 (0.36, 1.35) |
|  | 25-44 | 2.28 (1.36, 3.81) | 2.16 (1.25, 3.73) |
|  | 65+ | 4.65 (2.42, 8.94) | 5.34 (2.62, 10.87) |
| **Gender** | Male | Ref | Ref |
|  | Female | 0.44 (0.30, 0.66) | 0.42 (0.27, 0.63) |
| **Patient category** | Outpatient | Ref | Ref |
|  | Inpatient | 0.03 (0.02, 0.07) | 0.04 (0.02, 0.07) |
| **Rapid test done.** | No | Ref |  |
|  | Yes | 1.57 (0.94, 2.62) | - |
| **Days stayed at the hospital** | 0 to 1 day | Ref | Ref |
|  | 2-3 days | 0.62 (0.42, 0.93) | 0.53 (0.34, 0.80) |
|  | 4+ days | 0.54 (0.32, 0.91) | 0.48 (0.28, 0.85) |
| **Days delayed to hospital** | 0 days | Ref |  |
|  | 1+ Days | 1.23 (0.80, 1.90) | - |
| **Antibiotics were given.** | No | Ref |  |
|  | Yes | 0.97 (0.335, 2.68) | - |
| **Provinces (Location)** | Lusaka | Ref | Ref |
|  | Eastern | 1.07 (0.38, 2.96) | 0.82 (0.27, 2.56) |
| *95% Confidence Interval; Adjusting for Age and Sex* |  |  |  |

## Discussion

The study identifies significant determinants of mortality in cholera patients, specifically age, sex, inpatient status, and duration of hospital stay. These results are in line with findings across sub-Saharan Africa and will assist in planning targeted responses in future epidemics. Two distinct outbreaks occurred between January 2023 and July 2024, with the first outbreak, in early 2023, being minor and not very serious. This suggests that it was endemic and probably caused by localized contamination or inadequate sanitation. The second outbreak, which began in late 2023, spread quickly in January 2024 with nearly 300 daily admissions, signifying widespread transmission and health system strain as seen in most urban African outbreaks. The sudden increase and subsequent decline in the number of daily admissions suggests a successful public health response involving intensified disease surveillance, prompt case management, community sensitization, and WASH improvements ^[2,12]^

The study showed that most cases came from Lusaka Province, around densely populated areas with poor water and sanitation infrastructure, especially in informal settlements ^[13]^, while very few patients where from Eastern Province, with majority of patients there being generally older, suggesting different rural vs. urban transmission dynamics. Working-age adults, especially males, comprised the highest proportion of cases, a trend consistent with regional studies that found that mobile, working-age individuals faced a higher exposure risk to cholera ^[2,14]^.

Almost all patients were treated as inpatients, signaling either greater severity of illness or a policy strategy to manage all suspected cholera cases within treatment centers. Antibiotic use was overall low but significantly higher in Eastern Province, reflecting differences in protocols or resource access ^[4] .^ Hospital stays were brief, which was consistent with the presentation of an acute disease, such as cholera, and the rapid recovery following rehydration. Discharges within one day occurred frequently, especially in Lusaka, whereas Eastern Province patients were more likely to stay two to three days. The mean delay from symptom onset to healthcare arrival was short, though slightly longer in Eastern Province, where nearly half experienced delays known to be contributors to cholera mortality ^[15]^Overall, outcomes were mostly favorable, with majority of patients having recovered. However, mortality rates for both Lusaka and Eastern Province remained markedly higher than the WHO-recommended of less than 1%, indicating the need for strengthening earlier case identification, diagnosis and treatment ^[16]^

Bivariate analysis identified age, gender, outpatient status, rapid test result, and duration of hospital stay as significant mortality risk factors among cholera patients. Patients above 65 years had a higher mortality rate seconded by infants (<1 year). This is consistent with patterns in similar settings where the elderly and young children presented late or struggle with fluid loss ^[4]^. Men had higher mortality, aligning with Lusaka-based analyses ^[2]^

Outpatients faced dramatically higher mortality compared to inpatients, emphasizing the need for rapid identification of cases at community level, early fluid management and timely referral of severely ill patients to treatment centers for continued management. Comparable trends have been observed in Cameroon and DRC, where lack of inpatient access increased risk of death among cholera patients. Shorter hospital stays were also linked to worse outcomes, with patients who died having had shorter stays compared to survivors, reflecting possibly early deaths before adequate fluid administration; similar findings were reported during Malawi and Tanzania outbreaks ^[6,17]^

Multivariable analysis revealed significantly elevated death risk among patients aged 25 to 44 years (AOR 2.16) and ≥65 years (AOR 5.34) compared to those aged 45 to 64, consistent with prior regional studies highlighting older adults’ vulnerability, possibly due to comorbidities, delayed care-seeking, or physiologic susceptibility ^[12,18]^ Outpatient treatment remained an independent risk factor (inpatients AOR 0.04), illustrating the importance of adhering to case management protocols so that high-risk cases are promptly identified and managed in admission. Early deaths tied to shorter stays echo findings from sub-Saharan Africa, where rapid deterioration post-presentation is common ^[6, 17].^

These findings reinforce the necessity of early recognition and prompt referral for high-risk groups, especially older adults and infants, to inpatient care. They also emphasize the need for strong triage at outpatient facilities, community case management, and public health education on timely care-seeking. Long-term priorities remain strengthening WASH systems, surveillance, and vaccination campaigns, alongside sustained investments in health infrastructure. As cholera outbreaks grow more frequently with climate change and urbanization, evidence-based, tailored interventions are essential to protect vulnerable populations ^[2, 4, 19]^

## Acknowledgements

We gratefully acknowledge the Zambia National Public Health Institute (ZNPHI) and the Ministry of Health, Lusaka, Zambia, for their support and for providing access to the surveillance data used in this study.

## Data Availability

The data supporting the findings of this study are available from the Zambia National Public Health Institute (ZNPHI) (https://www.znphi.org.zm) and the Ministry of Health, Lusaka, Zambia (https://www.moh.gov.zm). Access to this data is restricted, as they were obtained under license for this study and are therefore not publicly available. Data may be obtained upon reasonable request and with prior permission from the relevant authorities.

